# Surveillance of NK cell subsets and cytokine profile in patients with Rocky Mountain Spotted Fever

**DOI:** 10.1101/2021.11.21.21266534

**Authors:** Carolina Maynez-Prieto, Jaime R. Adame-Gallegos, Moisés Ramírez-López, Estefanía Quintana-Mendías, Blanca E. Estrada-Aguirre, Maribel Baquera-Arteaga, Diana Manzanera-Ornelas, Enrique Guevara-Macías, Gerardo P. Espino-Solís

## Abstract

**Introduction:** The intracellular pathogen *Rickettsia rickettsii* causes tick-borne spotted fever (also called Rocky Mountain spotted fever (RMSF) and is increasingly recognized as an emerging cause of febrile illness in Mexico. However, little is known about the early immune responses to infection.

**Methodology:** Four RMSF pediatric patients on acute phase and eight healthy controls from Chihuahua, Mexico were recruited. The natural killer cell (NK) immunophenotype and the cytokine profile in peripheral blood were characterized by flow cytometry.

**Results:** There was a decrease in total NK (CD3^-^CD56^+^) and activation receptor (CD56^+^NKG2D^+^) in NK population in patients at day 3 after hospitalization, when compared to healthy controls. Regarding to the NK cytotoxic population (CD16^bright^CD56^dim^) there was an observed decrease in patients only, between day 3 and on hospital admission. Interleukin and chemokine levels measured were significantly increased in patients upon admission compared to controls (IL-6, IL-8, IL-10, IP-10, MCP-1 and MIG).

**Conclusions:** This study shows that circulating NK cells are numerically decreased, while cytokines induce a pro-inflammatory process in patients.

## 1 Introduction

Rocky Mountain spotted fever (RMSF) is a tick-borne disease caused by *Rickettsia rickettsii*, an obligate intracellular pathogen that belongs to α-protobacteria [1,2]. In general, tick-borne diseases have been overlooked as neglected tropical diseases despite their global distribution, although their inclusion in the WHO list has been suggested due to their relevance in Public Health [3]. In North America, RMSF is a reemerging infectious disease that primarily affects children living in poverty. In Mexico, in the states of Baja California and Sonora, 2,300 cases and 380 deaths were registered in 2016, and it is estimated that fatality rates are higher in children under 10 years of age [4]. In 2020 and 2021, 12 and 59 cases of RMSF were reported in the state of Chihuahua alone [5].

Rickettsia invades endothelial cells promoting their dissemination and causing vasculitis that stimulates cell signaling cascades, which leads to cytokine secretion, initiating infiltration into the vascular cell wall and perivascular space of macrophages, CD8^+^ T lymphocytes, CD4^+^ T lymphocytes and NK cells [6]. In human in vitro models, endothelial cells increase the expression and secretion of cytokines IL-1 (interleukin-1), IL-6 and TNFα (tumor necrosis factor α); chemokines such as CXCL8 / IL-8 (interleukin-8), CXCL10 / IP-10 (interferon gamma-induced protein 10), CCL2 / MCP-1 (monocyte chemoattractant protein 1), CXCL9 / MIG (interferon gamma-induced monokine) and CCL5 / RANTES that lead to the activation and recruitment of monocytes, NK cells (natural killers) and T lymphocytes to the site of infection leading to the potentiation of the inflammatory response and its elimination [7–11]. Monocytes, immature macrophages that circulate in the bloodstream, when infected promote the spread of the pathogen in the vascular endothelium, causing an increase in microvascular permeability leading to rickettsial vasculitis [12–14].

While NK cells, the main population of innate lymphocytes that coordinate early responses to bacterial infections, participate on the regulation of the immune response to rickettsia, particularly by preventing early endothelial injury that results in tissue damage [15,16]. In murine models, it is well established that NK cells are involved in early innate defense against rickettsial infection [17,18]. However, in humans, the host’s early immune responses have not been well characterized. In this study, we investigated serum cytokine and chemokine and NK responses in febrile children with acute *R. rickettsii* infection. Despite the limitations regarding the samples for this study, in 2020 there were 12 total cases of RMSF in Chihuahua and the results presented herein account for one third of all reported cases statewide.

## 2 Materials and methods

### 2.1 Study subjects and inclusion criteria

The study group included four patients with rickettsiosis (2 female and 2 males; mean age ± SD, 5 ± 5 years) from Children’s Hospital of Specialties of the state of Chihuahua and eight healthy controls matched by age and sex (4 female and 4 male; mean age ± SD, 5 ± 5 years). All samples were collected from September to November 2020. Everyone was punctured in the saphenous vein to obtain blood samples that were collected in EDTA anticoagulant tubes. The diagnoses of the patients required the detection of the gltA gene of *Rickettsia spp*. by polymerase chain reaction (PCR) using test kits, not having a history of autoimmune, cardiovascular, cerebral and / or osteoarticular disorders; secondary acute infectious processes, and ingestion of medications prior to the first sampling. All healthy controls were recruited from the state of Chihuahua, Mexico, which was the same area where the patients were infected. The controls must not have a history of tick bite prior to sampling; autoimmune, cardiovascular, cerebral and / or osteoarticular disorders; acute infectious processes, and ingestion of drugs prior to sampling.

### 2.2 Statement of ethics

The study protocol complied with the guidelines established in the Regulations of the General Health Law in matters of health research, the Declaration of Helsinki and the Good Clinical Practices issued by the National Bioethics Commission. It was also approved by the Research Ethics Committee of the Faculty of Medicine and Biomedical Sciences of the Autonomous University of Chihuahua with registration number CI-056-19.

### 2.3 Cytokine and Chemokine Profiles

Cytokine and chemokine levels was evaluated from sera samples obtained from patients with rickettsiosis and control donors. For this, the BD cytometric bead array (CBA) human inflammation kit (Catalog No. 551811), human Th1/Th2/Th17 kit (Catalog No. 560484) and human chemokine kit (Catalog No. 552990) were used following the manufacturer’s instructions. Briefly, reaction was made from a Master mix containing 5 μL of each antibody for the number of reactions to be done. Diluent solution was added to this mixture so that the volume per reaction was 50 μL of the Master mix. To this volume, 50 μL of the test sample and 25 μL of PE were added. The reaction was incubated for 2 h in the dark. Subsequently, 1 mL of wash buffer was added, and it was centrifuged for 5 min at 200× *g*. Finally, 350 μL of wash buffer was added and it was read on an Attune NxT cytometer in which 10,000 events were recorded at a flow of 100 μL/min. Results were analyzed with the FlowJo vX.0.7 program. A graph was generated comparing the size (FWD) with the complexity (SSC) of the spheres. In this way the regions of interest corresponding to the fluorescence of PE bound to the antibody were delimited and the fluorescence intensity corresponding to each cytokine was measured. With these values the concentration of each cytokine was calculated in picograms per milliliter (pg/mL).

### 2.4 NK cells Immunophenotype

100 µl whole blood were obtained from each sample and stained with the following monoclonal antibodies and reagents: anti-CD3 conjugated with Super Bright 600 (Clone: OKT3, # 63-0037-42), anti-CD16 conjugated with allophycocyanin-H7 (Clone: 3G8, # 560195), anti-CD56 conjugated with allophycocyanin (Clone: CMSSB, # 17-0567-42), anti-CD57 conjugated with Pacific Blue (Clone: TB01, # 12-0577-42), anti-NKG2D conjugated with Fluorescein Isothiocyanate (Clone: 1D11, # 11-5878-42), and anti-NKG2A conjugated with Brilliant ™ Blue 700 (Clone: 131411, No. 747926) (all from Thermo Fisher, Waltham, MA). The samples were incubated in the dark for 15 min, then 2 ml of lysis buffer (BD Solution BD FACS ™ 10X concentrated lysate, San Diego, CA) were added and incubated in the dark for 10 min. Subsequently, 2 ml of phosphate-buffered saline (PBS) were added and centrifuged at 1.800 rpm for 5 min to remove supernatant, 2 ml of PBS were added again, and they were centrifuged at 1,800 rpm for 5 min. After removing the supernatant, they were resuspended in 500 µl of PBS. Finally, the resuspended samples were acquired in the Attune NxT cytometer with four lasers (blue, red, violet and yellow) that allows us to read up to 14 parameters. Results were analyzed in FlowJo software where the Fluorescence minus one (FMO) controls and gate strategy to identify the populations was determined (version 10; BD science, San Diego, CA) (**Supplementary Figure 1**).

### 2.5 Statistical analysis

Data underwent a normality test using the Kruskall-Wallis test and Dunn’s test for multiple comparisons, P values less than 0.05 were considered statistically significant. GraphPad Prism version 9.3.1 software was used (GraphPad Software, San Diego, CA) for analysis and graphical work.

## 3 Results

At the time of admission, the patients reported clinical symptoms of average 5.5 days of evolution (2 to 9 days), characterized by frontal headache, fever, abdominal pain, and severe myalgia. Of the 4 patients, 3 were admitted to the general ward and discharged after 5-7 days, 1 of them was admitted to the Pediatric Intensive Care Unit (PICU) and was discharged after 13 total days at the hospital.

Levels of pro-inflammatory cytokines were evaluated (IL-1β, IL-6, IL-8, IL-10, IL-12p70 and TNFα) in sera from children with rickettsiosis in the acute phase of the disease. It was found a significant increase of IL-6, IL-8 and IL-10 in patients upon admission to hospital compared to controls. No variations were found in the concentrations of IL-1β, IL-12p70 and TNFα between patients and controls (**Figure 1**). When determining serum chemokine levels (IP-10, MCP-1, MIG, RANTES and IL-8), significant increase in IP-10, MCP-1, MIG and IL-8 was found in patients upon admission to hospital compared to controls. Whereas no variation was found in RANTES concentration between patients and controls **(Figure 2**).

**Figure 1.**
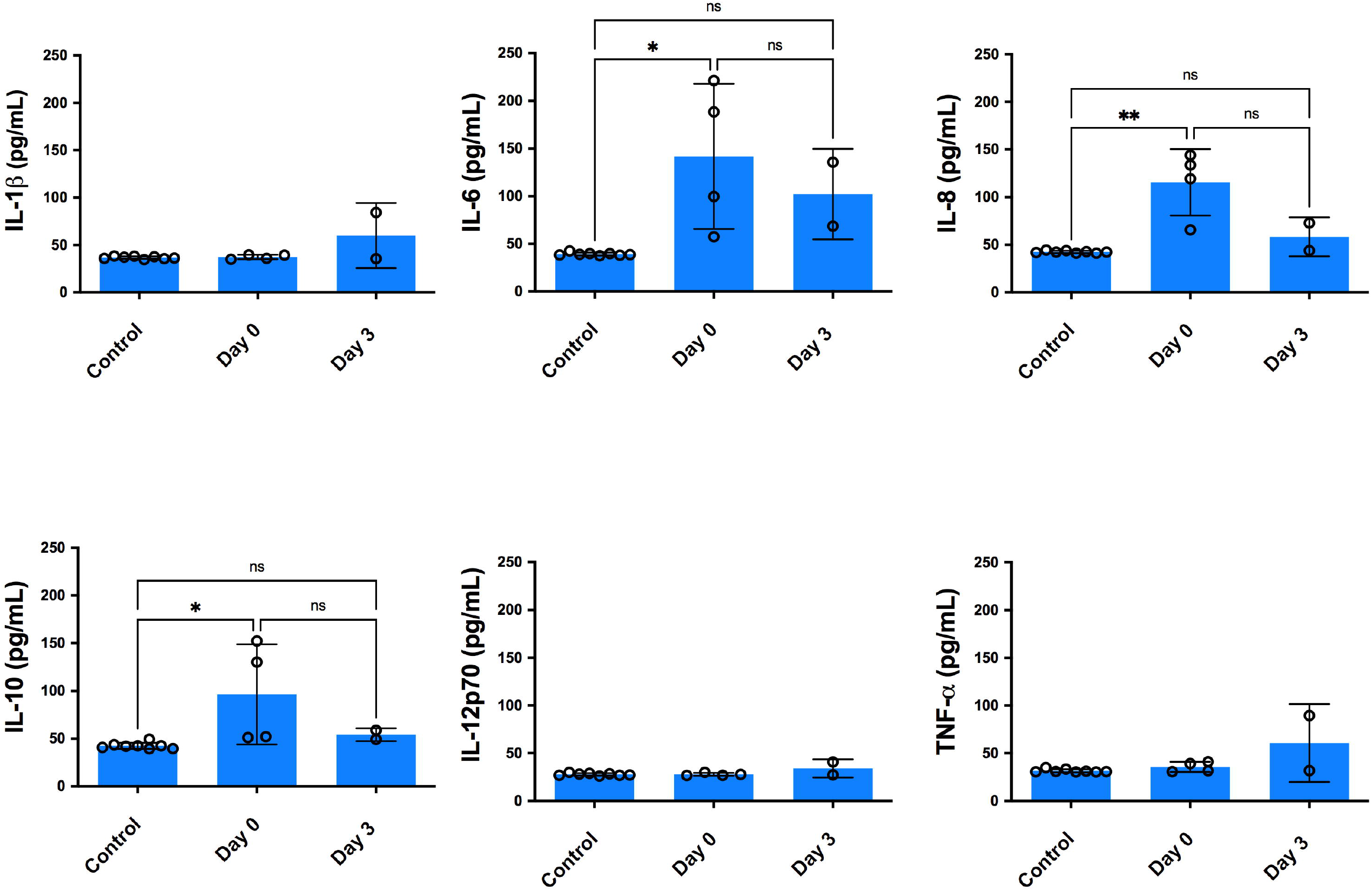
Proinflammatory cytokine levels in sera from children with rickettsiosis during the acute phase. Four sera from children infected with *Rickettsia rickettsii* and eight sera from healthy children were analyzed in parallel by the bead-based human inflammatory cytokine kit assay. Statistical analyzes were performed with the test of Kruskal-Wallis and Dunn’s subsequent multiple comparison test. Asterisks indicate differences statistically significant (* p <0.05, ** p <0.01).

**Figure 2.**
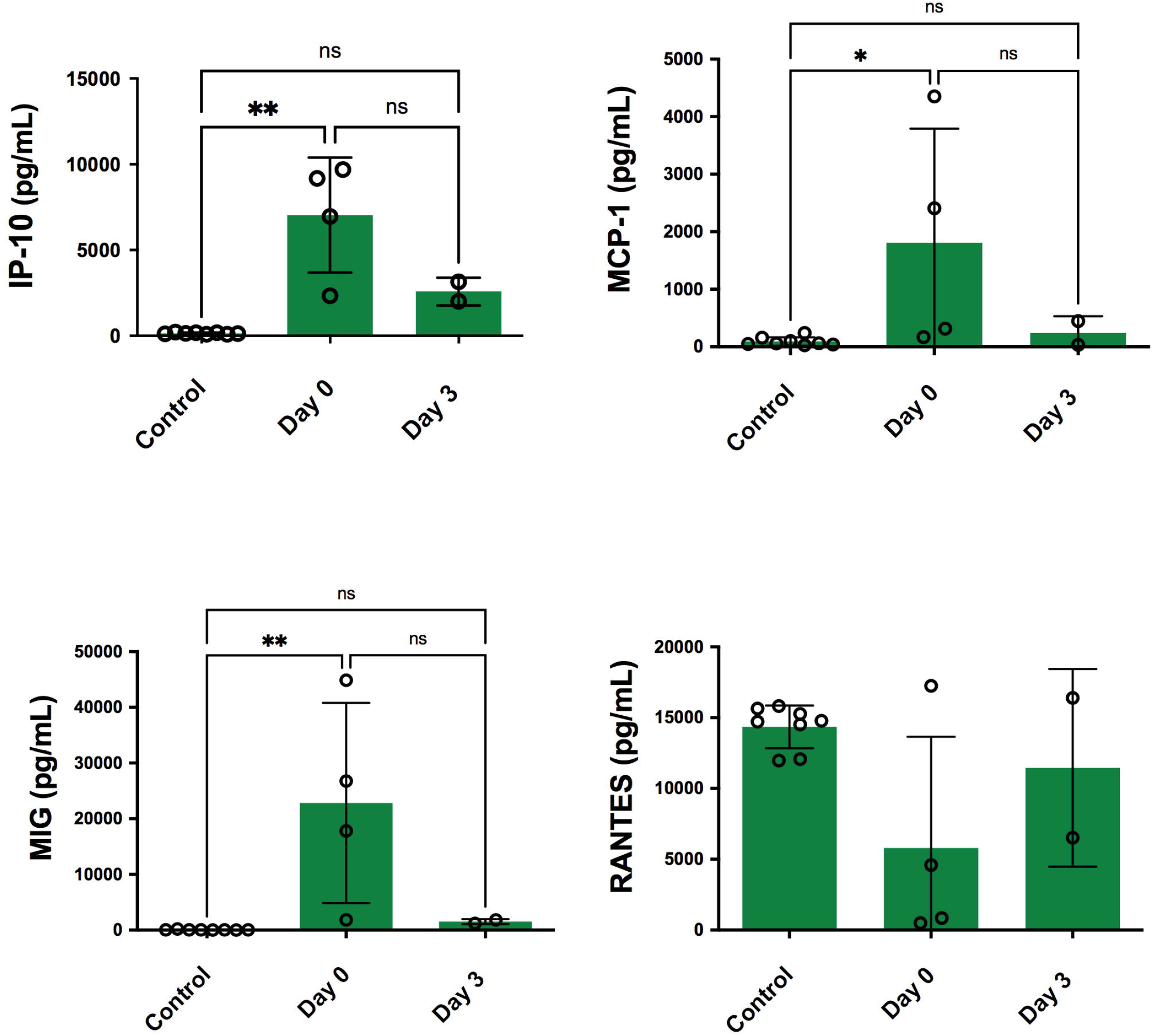
Chemokine levels in sera from patients with acute rickettsiosis. Sera obtained from children infected with *Rickettsia rickettsii* and eight sera from healthy children were analyzed in parallel by the bead-based human chemokine kit assay. Statistical analyzes were performed with the test of Kruskal-Wallis and Dunn’s subsequent multiple comparison test. Asterisks indicate differences statistically significant (* p <0.05, ** p <0.01).

Related to cytokines secreted during Th1 / Th2 / Th17 responses (IL-2, IL-4, IL-6, IL-10, IL-17A, IFN-γ and TNF-α), within the Th1 response, a significant increase in IFN-γ was found in patients upon admission to hospital compared to controls. No variation was found for TNFα and IL-2. In the Th2 response, a significant increase in IL-6 and IL-10 was found in patients upon admission to the hospital compared to controls, but not with IL-4, where there was no significant variation. In the Th17 response, there was no significant variation of IL-17A between patients and controls (**Figure 3**).

**Figure 3.**
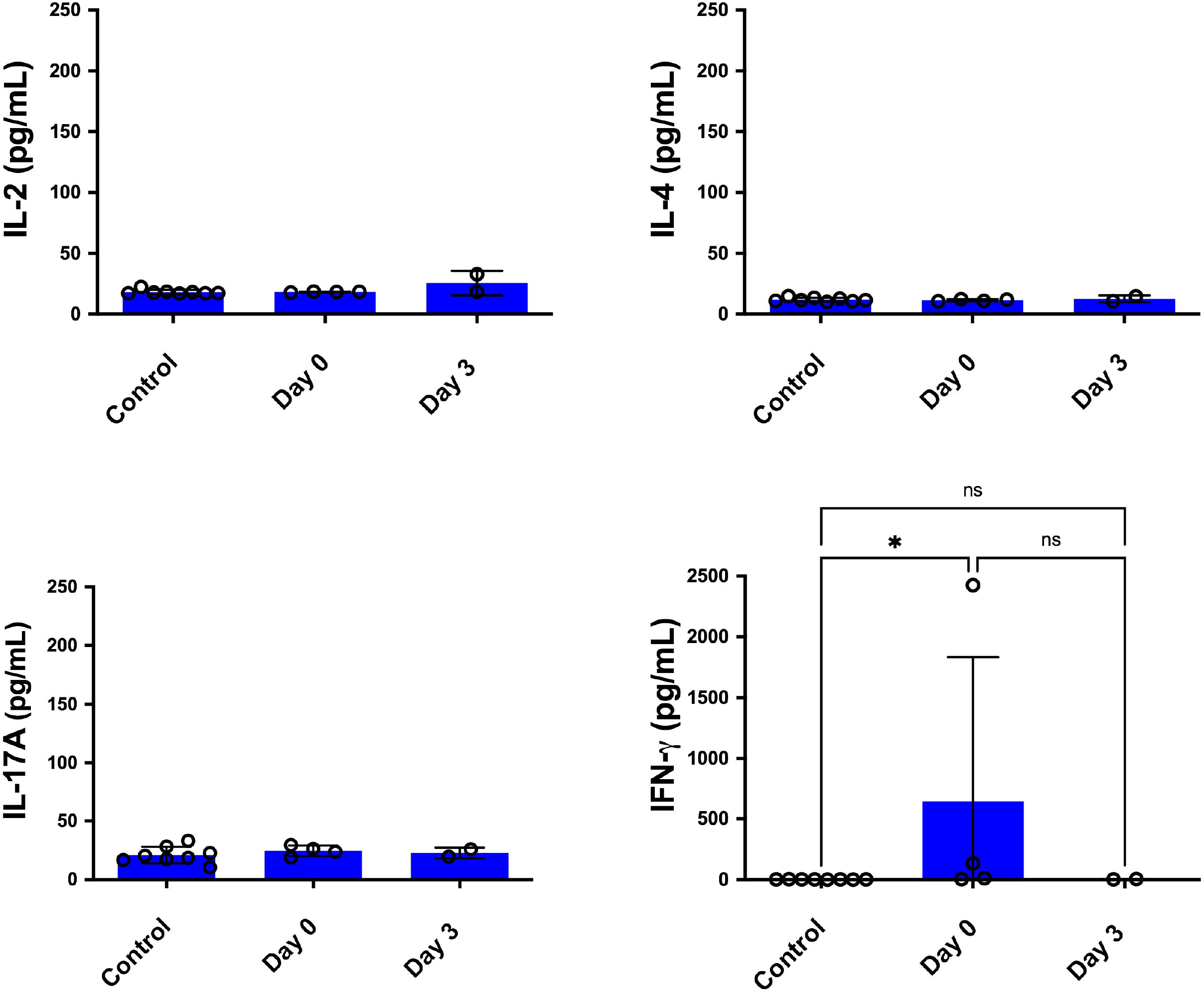
Levels of cytokines involved in Th1 / Th2 / Th17 responses in sera from children with rickettsiosis during the acute phase. Sera obtained from children infected with *Rickettsia rickettsii* and eight sera from healthy children were analyzed in parallel by the bead-based human Th1 / Th2 / Th17 cytokine kit assay. Statistical analyzes were performed with the test of Kruskal-Wallis and Dunn’s subsequent multiple comparison test. Asterisks indicate differences statistically significant (* p <0.05, ** p <0.01).

There was a decrease in total NK (CD3^-^CD56^+^) and activation receptor (CD56^+^NKG2D^+^) in NK population in patients at day 3 after hospitalization, when compared to healthy controls. Regarding to the NK cytotoxic population (CD16^bright^CD56^dim^) there was an observed decrease in patients only, between day 3 and on hospital admission. There were no significant differences in the percentage of NK effector population (CD16^dim^CD56^bright^), inhibition receptor (CD56^+^NKG2A^+^) and senescent cells CD56^+^CD57^+^) (**Figure 4)**.

**Figure 4.**
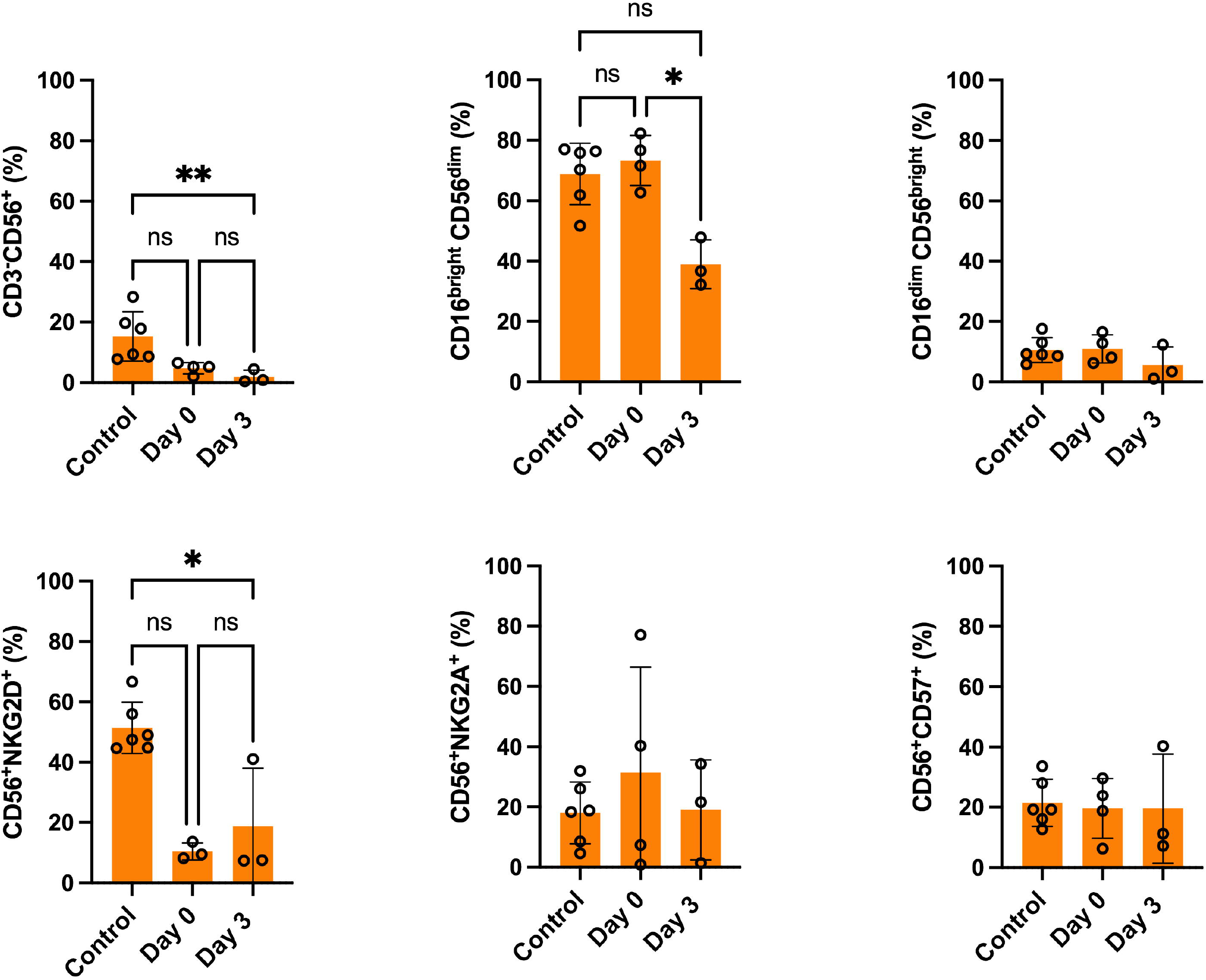
Percentage of NK cells (CD3^-^CD56 ^+^), effector NK cells (CD16^dim^ CD56^bright^), cells Cytotoxic NKs (CD16^bright^CD56^dim^), CD57-expressing NK cells (CD56^+^CD57^+^), NK cells expressing NKG2D (CD56^+^ NKG2D^+^) and NK cells expressing NKG2A (CD56^+^ NKG2A^+^) in blood obtained from children with rickettsiosis during the acute phase and healthy controls. Two analyzes were performed on parallel with four sera from children with rickettsiosis. The first analysis upon admission to the hospital and a second analysis 3 days after treatment, as well as whole blood from healthy children. Statistical analyzes were performed with the Kruskal-Wallis test and the subsequent Dunn multiple comparison test. Asterisks indicate differences statistically significant (p<0.01).

## 4 Discussion

Despite the small number of samples evaluated from patients with RMSF, we must take into consideration the ongoing COVID-19 situation at the time. Furthermore, only 12 cases were reported during this period in the state of Chihuahua. *In vivo* RMSF studies in humans evaluating circulating levels of inflammatory mediators and NK are scarce, probably due to the limited availability of patient samples. In the present pilot study, it was observed that in pediatric patients with RMSF it was characterized by a systemic increase in several pro-inflammatory cytokines such as IL-6 and IL-8. This increase has also been reported in other studies of patients with RMSF [9,19,20] reflecting the activation of monocytes / macrophages and endothelial cells infected by rickettsia, and attraction of T and NK lymphocytes [11,21].

Levels of IL-8, IP-10, MCP-1 and MIG showed a significantly high increase in patients upon admission compared to the controls. These chemokine levels are consistent with those reported in pediatric patients infected with *R. felis* during the acute phase, where IL-8, IP-10, and MCP-1 attract immune cells to infection sites and likely reflect initial host responses to *R. rickettsii* [9]. Plasma levels of IL-10, a potent inhibitor of TNFα and IL-6, were found to be significantly increased in patients upon admission to hospital. This result is consistent with previous studies in patients with other rickettsiosis (*R. africae* and *R. conorii*) where a significant increase in IL-10 was observed around day 12 after the onset of symptoms, possibly associated with the prevention of a disproportionate inflammatory response [19,22].

There was a decrease observed in circulating NK cells (CD3^-^CD56^+^) in the pediatric patients in this study, that has also been seen in patients with septic shock [23,24], and sepsis due to Gram positive bacteria such as *Streptococcus pneumoniae* and *Staphylococcus aureus*, and Gram-negative bacteria of the genus *Enterobacteria* and *Neisseria meningitidis*. In these cases, sepsis due to gram-positive bacteria the one that produced a greater decrease in NK cells circulating compared to sepsis due to gram-negative bacteria. This could indicate a persistent activation of NK cells during severe infection by gram-positive bacteria, the elimination of which normally requires highly organized host responses, whereas many gram-negative pathogens, as is the case in this study, could be effectively destroyed by complement and antibodies [25]. Furthermore, the observed reduction in the number of NK cells could reflect the activation and displacement of these cells to other tissues [16]. Additionally, previous studies in human acute viral infections such as influenza and hepatitis B infection also found a decrease in circulating NK cells, also related to the activation and displacement of these cells to other tissues [26,27]. However, other authors have reported an increase in NK cells in severe sepsis due to gram-negative bacteria and acute infection by scrub fever caused by *Orienta tsutsugamushi*, which could be related to increased hematopoiesis, recruitment and auto-proliferation of NK cells in response to cytokines and microbial stimulation [16,28,29].

When analyzing the immunophenotype, a decrease in cytotoxic NK cells (CD16^bright^CD56^dim^) was observed in the patients on the 3^rd^ day after admission. In turn, there was a significant decrease in the expression of NKG2D of the NK cells from the patients upon admission compared to the 3^rd^ day of their hospitalization, when compared to controls. This result could be related to the behavior observed during the infection by *O. tsutsugamushi* where, although no significant differences were observed in the proportion of CD56^bright^ / CD56^dim^ NK cell subsets between patients and controls, they exhibited an activated phenotype with higher expression levels of CD69 [29]. Based on this information, we suggest it should be considered to evaluate the expression of CD69, which participates in the induction of IFN-γ secretion [30]. This event in turn could be one of the main causes of the significant increase in IFN-γ plasma levels, also described in patients with murine typhus (*R. typhi*) and in a patient with infection by *R. slovaca*. The early response to IFN-γ activates intracellular bactericidal mechanisms to further control the spread of infection [20,21].

Despite the limitations of the panel used to detect NK cells in the present study, further experiments should be considered to elucidate the role of NK cells in the context of RMSF infections. For example, the expression of chemokine receptors associated with NK cells displacement to tissues or proliferation marker Ki67 for NK cells. These data could provide information on the health status of the pediatric patient and given that children represent a particular vulnerable group, it is even more important to be able to make an early diagnosis to treat them properly.

## Data Availability

All data produced in the present work are contained in the manuscript

## 5 Acknowledgments

Special thanks to Dra. Guadalupe Virginia Nevárez-Moorillón for her critical remarks. Authors also acknowledge, families and staff at Children’s Hospital of Specialties of the state of Chihuahua. Dr. Gumaro Barrios Gallegos, M.C. Gloria Edith Márquez Leos, Dr. Jorge Alain Carmona Sawatsky, and Dra. Leticia Ruiz González from the Health Department of the state of Chihuahua.

## 6 Author Contributions

GPES: Conceptualization, project administration, supervision, data analysis, review and editing of manuscript. CMP and JRAG: conceptualization, data analysis, original draft, review and editing of manuscript. EQM, BEEA: FACS analysis. MRL, MBA, DMO, EGM: sample collection and patient resources. All authors contributed to this article and approved the submitted version.

## Supplementary Material

**Supplementary Figure 1. Fluorescence minus one (FMO) controls and gate strategy and identification of human NK cells**. Whole blood was incubated with target cells for 15 minutes at 4°C, surface stained with fluorochrome-conjugated anti-CD3, anti-CD16, anti-CD56, anti-CD57, anti-NKG2Aand anti-NKG2D mAbs. Profiles demonstrate the gating strategy [side scatter (SSC) vs. forward scatter (FCS) and CD56 vs. CD3] for identification of NK cells (CD3^-^CD56^+^), effector NK cells (CD16^dim^CD56^bright^), cells Cytotoxic NKs (CD16^bright^CD56^dim^), CD57-expressing NK cells (CD56^+^CD57^+^), NK cells expressing NKG2D (CD56^+^ NKG2D^+^) and NK cells expressing NKG2A (CD56^+^ NKG2A^+^) cell populations. Profiles also show CD56 versus CD57, NKG2A and NKG2D staining on CD3^-^ CD56^+^ NK cells for one representative donor after incubation of whole blood sample with indicated target cells, analysis was carried out with FlowJo software.

