## Supplementary figures and images for "Surveillance of NK cell subsets and cytokine profile in patients with Rocky Mountain Spotted Fever"

### Supplemental Figure 1

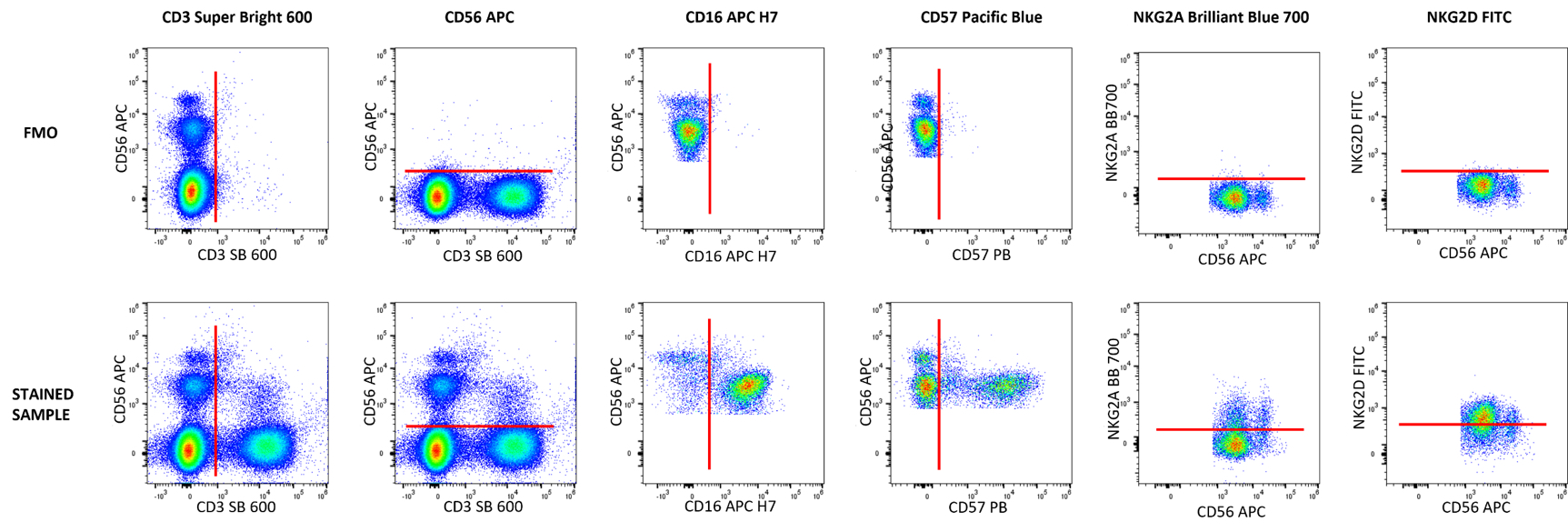
